# The iHealth-T2D study: rationale and design of a cluster randomised trial for prevention of Type 2 Diabetes amongst South Asians with central obesity and prediabetes

**DOI:** 10.1101/2020.11.12.20230433

**Authors:** Anuradhani Kasturiratne, Khadija I Khawaja, Sajjad Ahmad, Samreen Siddiqui, Khurram Shahzad, Lathika K Athauda, Ranil Jayawardena, Sara Mahmood, Mirthe Muilwijk, Tayyaba Batool, Saira Burney, Matthew Glover, Saranya Palaniswamy, Vodathi Bamunuarachchi, Manju Panda, Suren Madawanarachchi, Baldeesh Rai, Iqra Sattar, Wnurinham Silva, Swati Waghdhare, Marjo-Riitta Jarvelin, Ravindra P Rannan-Eliya, Heather M Gage, Irene GM van Valkengoed, Jonathan Valabhji, Gary S Frost, Marie Loh, Ananda R Wickremasinghe, Jaspal S Kooner, Prasad Katulanda, Sujeet Jha, John C Chambers

## Abstract

**Background:** People from South Asia are at increased risk of type 2 diabetes (T2D). There is an urgent need to develop approaches for prevention of T2D in South Asians, that are cost-effective, generalisable and scalable across settings.

**Hypothesis:** Compared to usual care, risk of T2D can be reduced amongst South Asians with central obesity or raised HbA1c, through a 12 month lifestyle modification programme delivered by community health workers.

**Design:** Cluster randomised clinical trial (1:1 allocation to Intervention or Usual care), carried out in India, Pakistan, Sri Lanka, and UK, with 30 sites per country (120 sites total). Target recruitment 3,600 (30 participants per site) with annual follow-up for three years.

**Entry criteria:** South Asian, men or women, age 40-70 years with i. Central obesity (waist circumference ≥100cm in India and Pakistan; ≥90cm in Sri Lanka) and / or ii. Prediabetes (HbA1c 6.0-6.4% inclusive). Exclusion criteria: known type 1 or 2 diabetes, normal or underweight (body mass index<22kg/m2); pregnant or planning pregnancy; unstable residence or planning to leave the area; serious illness.

**Endpoints:** The primary end point is new onset T2D at 3 years, defined as: i. HbA1c≥6.5% or ii. Physician diagnosis and on treatment for T2D. Secondary endpoints at 1 and 3 years are: i. Physical measures: waist circumference, weight and blood pressure; ii. Lifestyle measures: smoking status, alcohol intake, physical activity, dietary intake; iii. Biochemical measures: Fasting glucose, insulin and lipids (total and HDL cholesterol, triglycerides); and iv. Treatment compliance.

**Intervention:** Lifestyle intervention (60 sites) or Usual care (60 sites). Lifestyle intervention was delivered by a trained community health worker over 12 months (5 one-one session, 4 group sessions, 13 telephone sessions) with the goal of the participants achieving e a 7% reduction in body mass index and a 10 cm reduction in waist circumference through i. improved diet and ii. increased physical activity. Usual care comprised a single 30 minute session of lifestyle modification advice from the community health worker.

**Results:** We screened 33,212 people for inclusion into the study. We identified 10,930 people who met study entry criteria, amongst whom, 3,682 agreed to take part in the intervention. Study participants are 49.2% female and aged 52.8 (SD 8.2) years. Clinical characteristics are well balanced between Intervention and Usual care sites. More than 90% of follow-up visits are scheduled to be complete December 2020. Based on follow-up to end 2019, the observed incidence of T2D in the study population is in line with expectations (6.1% per annum).

**Conclusion:** The iHealth-T2D study will advance understanding of strategies for prevention of diabetes amongst South Asians, use approaches for screening and intervention that are adapted for low-resource settings. Our study will thus inform the implementation of strategies for improving the health and well-being of this major global ethnic group.

**IRB approval:** 16/WM/0171

**Trial registration:** EudraCT 2016-001350-18. Registered 14 April 2016 https://www.hra.nhs.uk/planning-and-improving-research/application-summaries/research-summaries/ihealth-t2d/ ; ClinicalTrials.gov NCT02949739. Registered 31 October 2016, https://clinicaltrials.gov/ct2/show/NCT02949739, First posted 31/10/2016.

**Funder:** European Commission (award 643774) and National Institute for Health Research (award 16/136/68)

## Background

South Asians, who represent one-quarter of the world’s population, are at high risk of Type 2 diabetes (T2D).^1^ India alone has ∼77 million people with T2D, the second highest number in the world.^2^ T2D prevalence is currently ∼5% in rural India, ∼11% in urban India.^3^ Similar patterns are observed among South Asians in Pakistan, Bangladesh, and Sri Lanka.^4^ The adverse consequences of diabetes are further magnified by the earlier age of onset of T2D, and by the low availability and financial barriers to obtaining high quality care in South Asia.^5^ South Asians living in Europe have a 2-4 times higher risk of T2D compared to Europeans, for reasons that remain to be determined.^5,6^ The high burden of T2D in South Asian thus represents a major public health challenge.

T2D is a preventable disorder. Amongst overweight and obese Europeans with impaired glucose tolerance, a programme of intensive lifestyle modification (increased physical activity, dietary change and weight loss) is associated with an ∼60% relative risk reduction in the incidence of T2D.^7^ The benefits are maintained for at least ten years after the intervention has stopped.^8^ Meta-analysis of published studies supports the potential for lifestyle intervention to reduce risk of T2D amongst South Asians.^9^ However, completed studies have been predominantly carried out in urban settings, and with under-representation of women, limiting the generalizability of findings. Furthermore, the trials have typically relied on identification of high-risk individuals using the oral glucose tolerance test, and on delivery of the lifestyle modification by trained healthcare teams, approaches that are resource intensive and difficult to scale-up. As a result, interventions for prevention of T2D are not routinely available to South Asian communities, especially in LMIC settings.

Delivery of health promotion for prevention of chronic disease has traditionally been the responsibility of physicians and allied health professionals. However, recent research supports the view that Community Health Workers (CHW) can make an effective contribution to prevention and early diagnosis of chronic disease, and deliver improved outcomes compared to usual care.^10^ Studies in rural India show that CHWs can provide health education and support management of hypertensive individuals.^11^ A comprehensive multicomponent intervention in Bangladesh, Pakistan, and Sri Lanka, showed that trained CHWs working in partnership with public health care systems, improved blood pressure control among adults with hypertension.^12^ Whether CHWs can deliver lifestyle modification for prevention of T2D in South Asians, that is effective, cost-effective and potentially scalable remains to be determined.

We therefore established the iHealth-T2D study, as a large-scale cluster-randomised clinical trial, to test the hypothesis that risk of T2D can be reduced amongst South Asians with central obesity or raised HbA1c, through a 12 month lifestyle modification programme delivered by CHWs.

## Preparatory work

### Development of a lifestyle intervention programme for delivery by CHWs

Our intervention programme was developed by a team with expertise in nutrition, dietetics, epidemiology and public health from the study centres in South Asia and the UK, and informed by the design of the Diabetes Prevention Programme.^7^ The primary objective was to achieve a 7% reduction in body mass index and a 10 cm reduction in waist circumference through i. **improved diet** and ii. increased physical activity. Improved diet comparised a 500 kcal energy reduction in energy intake for the overweight and obese. The profile of the diet was based on health eating guidelines which are common across non-communicable disease prevention programmes, and included attention to increasing fruit and vegetable consumption, decreasing sugar intake, and reducing alcohol consumption (where appropriate), portion sizes, and identifying cooking substitutions to reduce fat. **Increased physical activity** includes finding enjoyable physical activities to pursue regularly, and incorporating physical activity into daily routines. The target was to achieve 150 mins of moderate physical activity every week.

Core design elements of the programme included: 1) Goal-based behavioural intervention, with goals set in partnership with the individual on an ongoing basis, 2) Personalised support for the participant during delivery of the intervention, 3) Frequent contact to help participants achieve and maintain the weight and physical activity goals, 4) “Toolbox” strategies to tailor the intervention to the individual participant. Components specific to the current effort included: 1) Use of CHWs, supported by local experts for delivery of lifestyle intervention, 2) Cultural adaptation included development of meal plans and behaviour change advice suitable for South Asian communities, 3) Incorporating a family based approach to the intervention, intervention where one member of the patients family attended the education sessions, given the evidence for improved response to lifestyle advice when family members are included in the education,^13,14^ and 4) Making use of telephone and group sessions to bring peer support, to improve accessibility of the intervention, and to maximise efficiency of delivery. Relevant stakeholders (potential participants, CHWs, and other health care providers) were involved in the design and adaptation process.

The final lifestyle modification programme comprised 22 contact sessions over 12 months. The 22 sessions comprised five one-one meetings (duration up to 60 minutes), four group sessions (up to 90 minutes), and 13 telephone sessions (up to 15 minutes). The use of group and telephone contact sessions may improve participation, engagement and outcomes of intervention programmes compared to wholly face-face strategies.^15-17^ Session content and timing are summarised in **Table 1** and in the **Supplementary Online Materials**. The Intervention programme was supported by written materials for participants, translated into local languages. This participant Handbook contained educational information about the importance of obesity and diabetes, the potential benefits of lifestyle modification, and the key components of a healthy lifestyle (focused on improved and increased physical activity). The aim of the Handbook was to guide participants through the lifestyle modification sessions, and support self-monitoring of their lifestyle behaviours, goal setting, goal review and waist circumference.

**Table 1.**
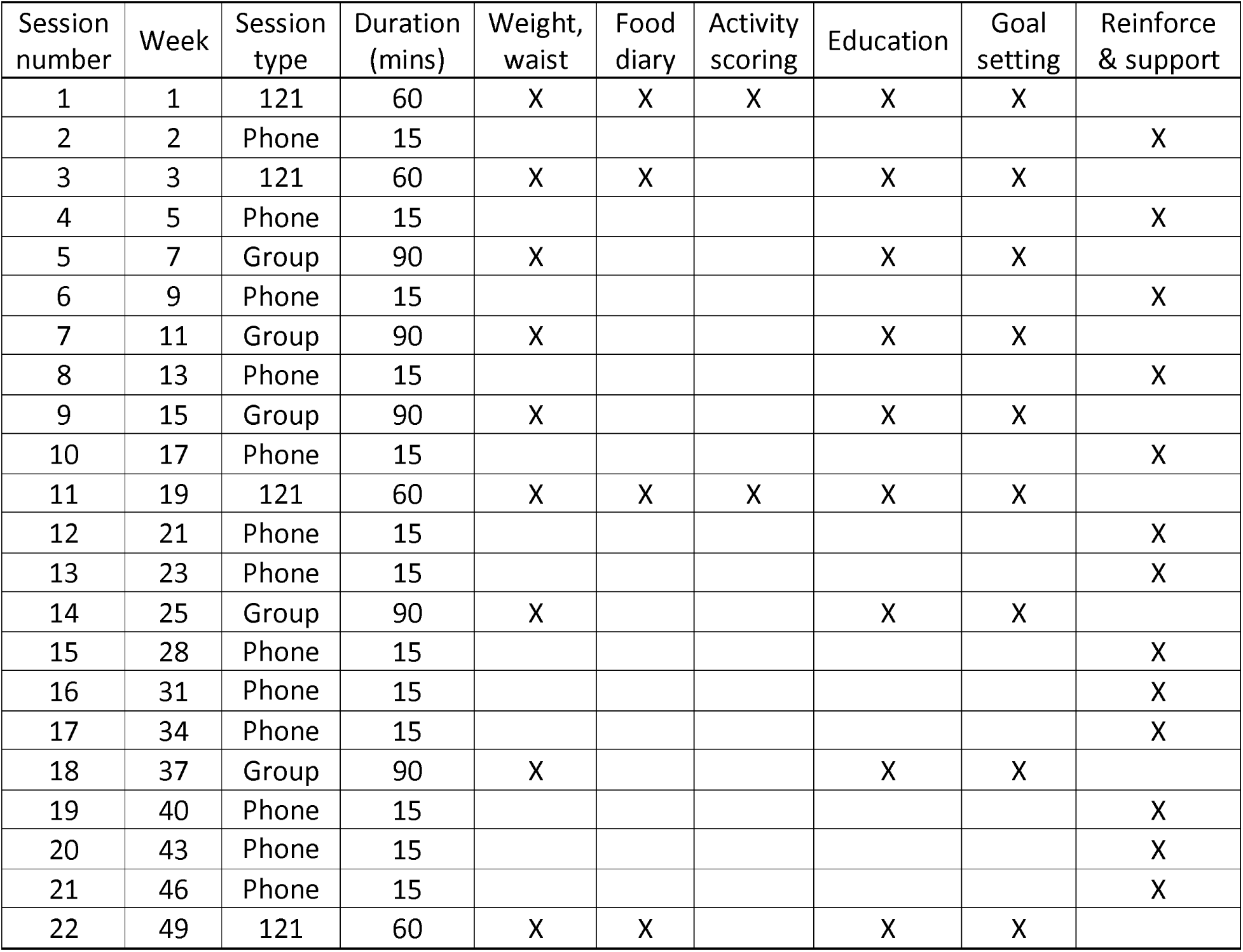
Timetable for the lifestyle intervention programme delivered by the community health workers. Overall there were 4 one to one sessions (121), five group sessions (Group), and 13 telephone support sessions (Phone).

### Identification and training of CHWs

CHWs were graduates with a biological science background, and were recruited from amongst the local community in which they worked, to ensure natural cultural awareness. Training of CHWs was provided by local experts in diabetes, nutrition and exercise, according to the principles and practices described in the intervention protocols. Training covered theoretical knowledge as well as practical hands on learning in communication and delivery of the intervention, and lasted 2-3 weeks. The study handbook summarised the protocols to be followed, and provided toolkits for promoting healthy diet and physical activity. All trainee CHWs were assessed for knowledge, skills and attitude by the trainers prior to commencing intervention. Each CHW was expected to initiate intervention for 80-100 new participants per year, followed by delivery of the complete programme over the 12 months.

### Identification of South Asians at increased risk of diabetes

The glucose tolerance test has been the mainstay for identification of future risk for T2D. However, this is a resource intensive approach, that is not well suited for implementation at population scale. To address this limitation, we used our longitudinal studies of South Asian populations to identify non-laboratory and laboratory markers of risk that may be better suited to identification of susceptible individuals from the general population. We compared a range of clinically relevant, routinely available measures for prediction of new onset T2D, amongst the 17,000 South Asian men and women aged 35 to 75 years, under long term follow-up in the London Life Science Prospective Population (LOLIPOP) study.^6^ Our results show that waist circumference is a strong predictor of incident T2D in South Asians, that offers better discrimination than body mass index or waist-hip ratio (**Table 2**). Waist circumference is a simple, readily available clinical measure of adiposity that is well-suited for use as a tool for community-wide risk stratification, especially in low-middle income countries. Our results also identify HbA1c as a highly predictive biochemical tool for identification of high-risk South Asians,^6^ that achieves similar discrimination for T2D to fasting glucose (**Table 2**). HbA1c has additional the advantage of being a non-fasting assay, which is simpler and cheaper to administer than an oral glucose tolerance test. HbA1c is potentially well suited for community-wide and opportunistic screening. These observations provide the rationale to investigate the clinical utility of HbA1c and waist circumference for identification of South Asians at high risk of T2D, in the present study.

**Table 2.** Risk factors for incident T2D (5 years) amongst South Asians in the LOLIPOP study.^6^

| Risk factor | Prevalence | T2D incidence | Sensitivity | RR for T2D |
| --- | --- | --- | --- | --- |
| *Biochemical* |  |  |  |  |
| Fasting glucose $\geq 6.0$mmol/L (WHO) | 5.0% | 55.6% | 19.3% | 6.66 (4.89-9.07) |
| HbA1c $\geq 6.0$% (WHO) | 16.3% | 68.4% | 35.0% | 13.9 (10.4-18.6) |
| *Anthropometric* |  |  |  |  |
| Body mass index $>28$ kg/m<sup>2</sup> (WHO) | 17.0% | 30.1% | 32.6% | 2.35 (1.96-2.82) |
| Waist-hip ratio (M$>1.0$, F$>0.9$) | 28.0% | 27.1% | 46.2% | 2.21 (1.88-2.60) |
| Waist: M or F $\geq 100$cm | 28.9% | 31.3% | 49.6% | 2.56 (2.21-3.06) |

## Methods

### Study design

iHealth-T2D is a multi-centre, cluster-randomised clinical trial to evaluate intensive lifestyle modification delivered by CHWs for prevention of T2D, compared to usual care. Study entry criteria were: South Asian, men or women, age 40-70 years with i. Central obesity (waist circumference ≥100cm in India and Pakistan; ≥90cm in Sri Lanka) and / or ii. Prediabetes (HbA1c 6.0-6.4% inclusive). Exclusion criteria were: Known type 1 or 2 diabetes, normal or underweight (body mass index<22kg/m2); pregnant or planning pregnancy; unstable residence or planning to leave the area; serious illness. We aimed to recruit 3,600 participants from 120 locations across 4 countries (India, Pakistan, Sri Lanka and the UK) comprising 900 participants and 30 sites per country. Participants were cluster randomised based on study site to receive either intensive lifestyle modification (N=60 sites) or usual care (N=60 sites). Participants will be followed at 12, 24 and 36 months after enrolment. The primary endpoint is the incidence of T2D at three years. The study was approved by an Institutional Review Board (IRB) in each participating country (ref 16/WM/0171), and was registered on the EudraCT database (2016-001350-18). Primary funding for the study was from the European Commission (award 643774). Imperial College London acted as the sponsor for the study. The complete study protocol and study materials are available on our website: www.ihealth-t2d.org.

### Study locations

Participants were recruited from 120 sites divided equally between India, Pakistan, Sri Lanka and UK (i.e. 30 recruitment sites per country). On the Indian subcontinent, study sites were distributed across a range of socio-economic and geographic settings, to increase the generalizability of the findings. Locations were identified based on the knowledge and advice of local experts and available administrative data for the region. In the UK, the study sites comprised GP surgeries in West London (London boroughs of Ealing, Harrow and Hounslow), selected as those serving populations with a high proportion of South Asians.

In India, our 30 study sites were distributed around five regional hubs comprising New Delhi (Delhi), and cities in four North Indian states: Gurugram (Haryana), Vaishali (Ghaziabad, Uttar Pradesh), Mohali (Punjab) and Muzaffarpur (Bihar). Delhi, officially known as the National Capital Territory (NCT) is a city and a union territory of India which includes New Delhi, the capital of India. There were six study sites (three rural and three urban or semi-urban) for each regional hub.

In Pakistan, the study area was the metropolitan district of Lahore, the second most populous city of Pakistan. It has an estimated population of approx.10 million with urban: rural population in 2:1 ratio. The city is divided into 10 basic administrative divisions (Towns) that are further divided into smaller units termed as Union Councils (urban) and Mouzas (rural/village). Our 30 sites represented all 10 towns with an average of 3 sites per town. The boundaries of each site were based on the respective Union Council / Mouzas defined by the main roads encompassing the area. There were 20 urban and 10 rural sites. The urban sites were congested; thus smaller and represented one union council. Rural sites had less population density, and thus were large and represented 2 or more mouzas or villages. The 30 study sites were co-ordinated through three hubs: Services Institute of Medical Sciences (SIMS), Punjab Institute of Cardiology (PIC) and Sharif Hospital.

In Sri Lanka the study was conducted in Colombo and Gampaha districts, the two most urban and populous districts of the country. Colombo district has 13 administrative divisions. We selected two (2) of these divisions, each representing predominantly urban and predominantly rural populations. Eight (n=8) clusters were selected from the urban setting and seven (7) clusters were selected from the rural setting. A cluster was defined as a Grama Niladhari division. Gampaha district has 13 administrative divisions. Only seven (7) of these divisions have urban populations. We sampled one urban cluster (n=7) from each of the divisions with urban populations. One cluster each was selected from the other six divisions with only rural populations (n=6). Two additional rural clusters were selected from the two most populous divisions that have both urban and rural populations (n=2). A cluster was defined as a Grama Niladhari division with a Primary Care Hospital (Primary Medical Care Unit/ Divisional Hospital) or a Maternal and Child Health clinic where screening can be conducted.

The 120 research sites were cluster randomised to Intervention or Usual care (1:1 allocation). Randomisation was using computer generated random numbers, and as stratified by country to ensure 15 intervention and 15 usual care sites per country. Randomisation was carried out before recruitment started.

### Recruitment to the study

We invited South Asian men and women aged 40-70 years and without known diabetes to be considered for the study. Recruitment was a two-stage process: i. an initial Screening Visit to assess eligibility for the intervention trial; ii. an Enrolment visit, to confirm eligibility and to obtain consent for inclusion in the intervention phase of the trial.

In South Asia, our primary recruitment strategy comprised an open invitation to individuals living in the community at study sites, to attend for an iHealth-T2D Screening visit. We first discussed the study with the relevant administrative and health authorities to engage their support and permission to operate. We held discussions with community leaders and open meetings to identify suitable approaches for engaging local communities in the project. We distributed knowledge of the project and invited people to attend for screening through trusted sources of health information (e.g. health centres, physicians and health care providers, grass root level non-physician health workers, accredited social health activists and volunteer groups). People were encouraged to discuss the project with neighbours and friends to help widen engagement. The screening health assessment was held in local community health centres easily accessible to the target community.

In the UK, our primary strategy was postal invitation. We sent letters by post, to men and women aged 40-70 years, registered to the practice lists of collaborating general practitioners and who were recorded to be of South Asian ethnicity, and not known to have diabetes. We sent 14,564 invitations, from which 3,031 attended a screening visit (22%). From these, 408 attended enrolment and 240 were recruited to trial (i.e., 1.6% of people invited were ultimately recruited to the intervention phase). Screening was carried out in local GP surgeries, and subsequent visits at a dedicated research clinic based at Ealing Hospital. As a secondary strategy in the UK, we also took advantage of a database comprising the results of cardiovascular health assessments recently carried out amongst 9,699 South Asians in the study sites, as part of the LOLIPOP study (2010-2015). These baseline assessments were done using methods identical to those of iHealth-T2D Screening visit, and are thus directly comparable. This approach enabled identification of South Asians meeting the study entry criteria based on HbA1c or waist circumference, who were then invited directly to a study Enrolment visit using a single letter of invitation (2,872 invitations sent; 1,026 attended; 36% response rate). From these, 579 were recruited to trial (i.e., 20.2% of people invited). This secondary strategy also enables the evaluation of the cost-effectiveness for recruitment to lifestyle intervention based on existing, routinely collected healthcare data, compared to using systematic population-based screening strategies.

### Screening and enrolment visits

Potential participants were invited to attend an initial Screening Visit to assess them for eligibility (Table 3). This comprised: i. Questionnaire (socio-demographic details, medical and drug history with emphasis on diabetes and cardiovascular disease, smoking and alcohol consumption); ii. Physical measurements (waist circumference, height, weight and blood pressure), and iii. Fasting blood samples (overnight) for measurement of HbA1c (study entry criterion), and for storage to enable epidemiological research into biomarkers for metabolic and cardiovascular health. People reaching the study entry criteria based on the initial screening assessment were invited to attend for an Enrolment visit. The data collected at Enrolment is the baseline data against which subsequent health outcomes will be assessed. In addition to repeating the measures collected in the screening visit, participants also completed the Global Physical Activity Questionnaire (GPAQ) and Health-related quality of life questionnaire (EQ-5D-5L). South Asians who continued to meet the study eligibility criteria were invited to enrol into the intervention phase of the study (Figure 1).

**Table 3.** Schedule visits for the iHealth-T2D study. GPAQ: generalised physical activity questionnaire. Quality of life: EQ-5D-5L.

|  | Screening | Enrolment | Follow-up 1 | Follow-up 2 | Follow-up 3 |
| --- | --- | --- | --- | --- | --- |
| Time | -30 days | 0 | 12 months | 24 months | 36 months |
| Consent | X | X |  |  |  |
| Demographics | X |  |  |  |  |
| Alcohol intake | X | X | X | X | X |
| Tobacco intake | X | X | X | X | X |
| Medications | X | X | X | X | X |
| Medical history | X | X | X | X | X |
| EQ-5D-5L |  | X | X | X | X |
| GPAQ |  | X | X | X | X |
| 24 hour Food diary |  | X | X | X | X |
| Height | X |  |  |  |  |
| Weight, waist, Hip | X | X | X | X | X |
| Blood pressure | X | X | X | X | X |
| HbA1c | X | X | X | X | X |
| Biological samples | X | X | X | X | X |

**Figure 1.**
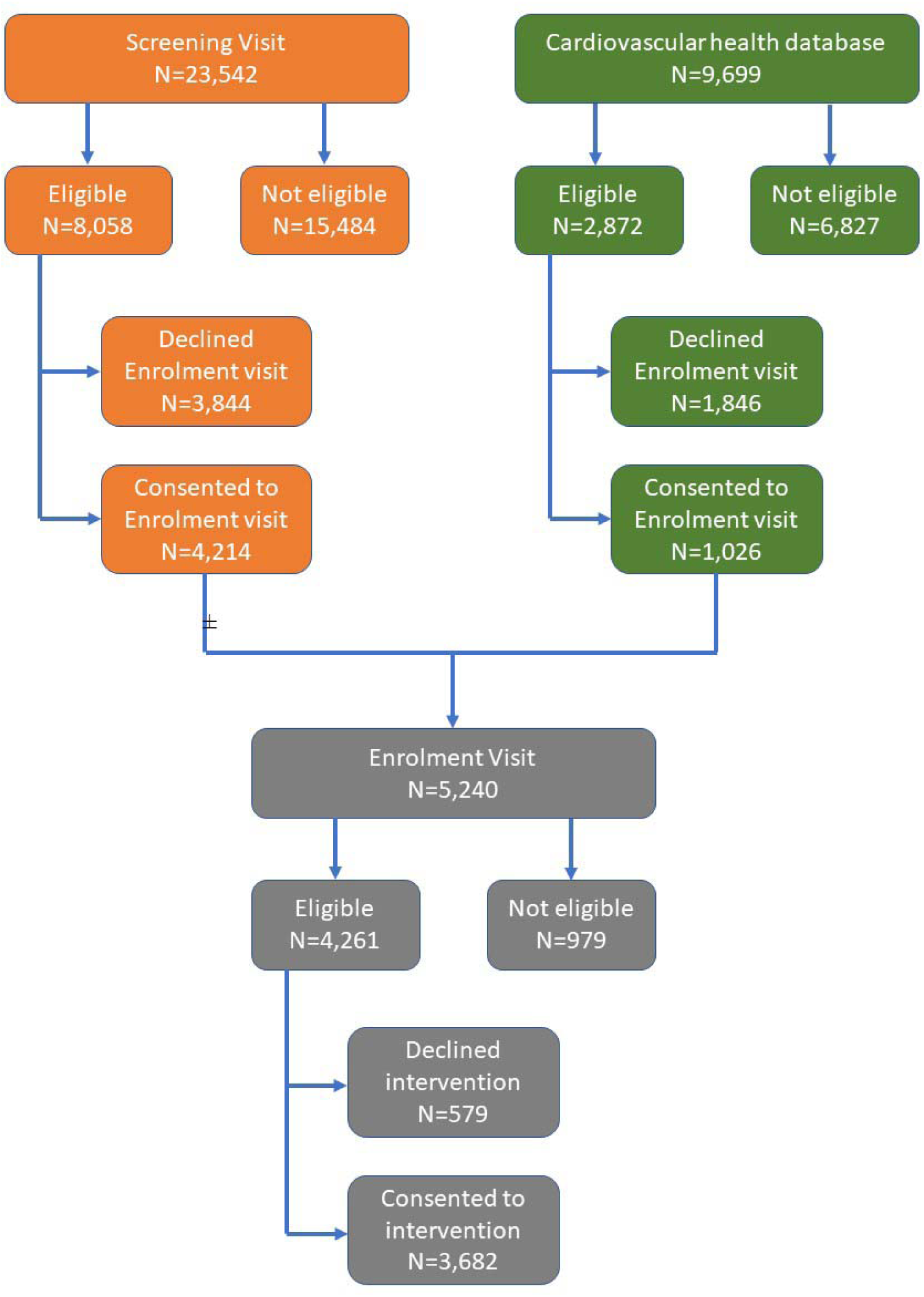
Overview of recruitment to the study.

All measures were carried out according to standardised protocols. Waist circumference was measured at the midpoint between the lower margin of the ribs and the top of the iliac crest. Hip circumference was measured at the level of the greater trochanters. Waist and hip measurements were measured three times and the average calculated. Weight was measured in light clothing using portable digital scales accurate to 0.1kg, and height using a portable stadiometer. Blood pressure was measured in the seated position using Omron digital devices. Three measurements were obtained from each participant, one minute apart. All people attending for a study visit were given a printed report of their results, along with written guidance on their interpretation. HbA1c was measured using Biorad D10 or Variant II turbo assays. Assays were performed in local laboratories that participate in external quality control, and meet the standards of the International Federation of Clinical Chemists. All participants with newly detected medical conditions were referred to the local health service.

### Study intervention

At the 60 study sites randomised to intervention, lifestyle modification was provided by the trained study CHW to the index cases, over 12 months (22 contact sessions), using the approaches set out in the study protocol. Family members of the index case living in the same household were encouraged to take part in lifestyle modification. Participation of family members was optional.

At the 60 study sites randomised to Usual care, participants received a single episode of brief lifestyle intervention for diabetes prevention, supported by written material. This session lasted 30-60 minutes and was delivered by the CHW.

### Follow-up visits

All participants enrolled to the study were asked to attend follow-up visits at 12, 24, and 36 months after recruitment. Follow-up assessments were conducted by a member of the research team, and not by the CHW providing the intervention. At each follow-up visit study participants completed a structured assessment similar to the enrolment evaluation. This included: i. Follow-up questionnaire; ii. Physical measurements (waist circumference, weight and blood pressure); iii. GPAQ and EQ-5D-5L; and iv. Fasting venous blood sample for measurement of HbA1c and serum biomarkers. Protocols and equipment were identical to those used for the baseline evaluation.

### Blinding and data handling

As the trial involves active intervention on both arms, it was not possible for participants and intervention staff to be blinded. However, outcome assessors and data analysts were kept blind to the allocation. The trial established procedures to maintain separation between staff that take outcome measurements and staff that deliver the intervention. Staff members who obtained outcome measurements were not directly informed of the group assignment. Intervention staff and dieticians who delivered the intervention did not take outcome measurements during follow-up. All investigators, staff, and participants were kept masked to outcome measurements and trial results.

### Study administration

The Steering Committee provided overall supervision of trial conduct. Day-to-day management of the study was co-ordinated through the Project Management Team, based at Imperial College London working in partnership with the project co-ordinators at each of the study centres. The Project Management Team was responsible for day-day administrative and scientific management of the project, including study planning, organisation of meetings, delivery of the annual reports, trial master files and compliance. Further details about the project are available on the project web-site (www.ihealth-t2d.eu).

## Statistical analysis and data management

All analyses will be carried out according to our published statistical analysis plan (SAP).^18^ Full details of the statistical approach are described in the SAP, and are briefly summarized here.

### Study endpoints

The primary end point is new onset T2D in the index case, defined as: i. HbA1c≥6.5% or ii. Physician diagnosis and on treatment for T2D. Secondary endpoints are: i. Physical measures: waist circumference, weight and blood pressure; ii. Lifestyle measures: smoking status, alcohol intake, physical activity, dietary intake; iii. Biochemical measures: Fasting glucose, insulin and lipids (total and HDL cholesterol, triglycerides); and iv. Treatment compliance.

### Sample size and power calculations

The study aimed to include 3,600 participants. Assuming that even rates for T2D for usual care are 6.8% per year, the study has 81% power to identify at *p*<0.05 a reduction in T2D incidence of 35%, after a follow-up period of three years with an estimated drop-out rate of 10% within the total population ^9^.

### Data Analysis

The primary analyses, conducted after three years of follow-up, will investigate whether intensive lifestyle modification reduces the risk of new onset T2D (primary endpoint) compared to usual care amongst South Asians with: i. Central obesity (N∼2,700 participants); ii. Raised HbA1c (N∼900 participants) and iii. Overall (central and / or prediabetes, all ∼3,600 participants). The analyses will be performed according to the intention to treat principle with data from all participants enrolled in the study.^19^ Random effects logistic regression will be used to estimate odds ratios for incidence of T2D and 95% CI. Secondary outcomes will be evaluated both after one and three years of follow-up, which will allow for comparisons of both short and long term effects of the lifestyle intervention. Subgroup analyses amongst participants included in the study based on HbA1c and/or waist circumference will be performed. The differences between the two treatment arms will be estimated with a multilevel linear mixed-effects regression model.

## Health economic analysis

The primary aim of the health economic analysis is to determine the cost-effectiveness of lifestyle modification vs usual care for prevention of T2D amongst i. South Asians on the Indian subcontinent and ii. South Asians in Europe. The health economic analyses will include pre-specified subgroup analyses of cost-effectiveness between: i. South Asians identified to be at high risk of T2D based on central obesity vs raised HbA1c≥6.0%; ii. Low-middle-income (Indian subcontinent) vs high-income (UK) regions; and iii. Socio-demographic: sex, socioeconomic classes, and across ages. The results of the cost-effectiveness analyses will be used to describe the potential clinical and financial implications of implementing the approaches into standard practice in the local and national health economies. Effectiveness will be expressed in Quality adjusted Life Years (QALYs), using responses to the EQ-5D-5L, as well as T2D cases prevented. Cost-effectiveness analyses will be performed based on the follow-up period, estimating incremental costs and QALYs using multi-level linear regression models in line with statistical analysis of other study endpoints, as well as using parametric extrapolation techniques to evaluate the intervention over longer time horizons. The estimates will enable assessment of the scalability of an intensive lifestyle modification intervention delivered by CHWs for prevention of T2D, using central obesity and / or HbA1c for screening. The analyses of the costs of scaling-up will take into account available and future resource envelopes in the South Asian countries, and also current and future pricing for assays such as HbA1c, to understand current and future feasibility.

## Results

Participants were recruited between 15/06/2016 and 05/03/2019. **Figure 1** shows the trial profile. In total, 33,212 people were screened for inclusion into the study. There were 23,542 people who attended an iHealth-T2D Screening Visit (**Table 4**). There were 10,930 people who met study entry criteria after the Screening Visit, and who were invited to attend for the Enrolment Visit. Amongst these, 5,240 people attended, of whom 4,261 people (81.3%) continued to meet study inclusion criteria. The primary reasons for participants not being eligible at the enrolment assessment were: waist circumference initially raised, but below threshold on repeat (N=251), HbA1c initially 6.0-6.4%, but <6.0% on repeat (N=375), repeat HbA1c ≥6.5% (N=200) or now diagnosed T2D (N=42), repeat BMI <22kg/m^2^ (N=34), and incomplete data (N=41).

**Table 4.** Characteristics of the 23,543 participants attending a Screening visit

|  | **India** | **Pakistan** | **Sri Lanka** | **United Kingdom** |
| --- | --- | --- | --- | --- |
| N | 6913 | 6962 | 6535 | 3132 |
| Female gender (%) | 37.7% | 47.6% | 68.2% | 53.1% |
| Age (years) | 51.6 (8.8) | 51.8 (8.6) | 53.2 (8.7) | 54.6 (8.8) |
| Current smoking (%) | 6.2% | 9.3% | 7.3% | 7.1% |
| Skilled worker (%) | 58.0% | 25.6% | 28.3% | 51.9% |
| Married (%) | 96.2% | 94.0% | 84.8% | 84.2% |
| Body Mass Index (kg/m<sup>2</sup>) | 27.5 (4.6) | 28.7 (5.7) | 25.3 (4.4) | 27.2 (4.5) |
| Systolic BP (mmHg) | 133.8 (18.1) | 130.4 (20.4) | 134.2 (19.8) | 127.8 (17.0) |
| Diastolic BP (mmHg) | 81.9 (10.6) | 80.8 (12.1) | 78.6 (11.0) | 79.3 (10.1) |
| Waist (cm) | 98.1 (11.2) | 97.9 (12.1) | 87.8 (10.9) | 93.1 (11.9) |
| Central obesity (%) | 46.9% | 43.9% | 26.4% | 27.1% |
| HbA1c (%) | 6.1 (1.4) | 6.3 (1.5) | 6.0 (1.2) | 5.6 (0.5) |
| Prediabetes (%) | 19.2% | 31.2% | 23.5% | 18.8% |
| HbA1c≥6.5% (%) | 21.8% | 24.2% | 14.4% | 2.7% |

Of these 4,261 people meeting study entry criteria after completion of the enrolment visit, 3,682 (86.4%) agreed to take part in the intervention phase (**Table 5**). There were no differences in gender between those who agreed or declined to take part in intervention (agreed: male 86.6%, female 84.8%, P=0.09). There were also no material differences in age (53.6±8.7 vs 55.0±8.7 yrs), body mass index (30.3±4.5 vs 30.5±4.9 kg/m^2^), waist (104.0±9.3 vs 103.0±10.7 cm) or HbA1c (5.76±0.43 vs 5.77±0.47 %) amongst those agreed to take part in the intervention compared with those who declined, respectively (all P>0.05).

**Table 5.** Characteristics of the 3,682 participants starting intervention

|  | **India** | **Pakistan** | **Sri Lanka** | **United Kingdom** |
| --- | --- | --- | --- | --- |
| N | 909 | 1070 | 884 | 819 |
| Female gender (%) | 46.8% | 48.5% | 67.9% | 32.5% |
| Age (years) | 50.5 (8.0) | 51.7 (7.8) | 53.4 (7.8) | 59.3 (8.5) |
| Current smoking (%) | 4.1% | 7.6% | 5.2% | 5.6% |
| Body Mass Index (kg/m<sup>2</sup>) | 30.2 (4.2) | 31.2 (5.2) | 28.9 (3.9) | 30.7 (4.2) |
| Systolic BP (mmHg) | 131.4 (14.8) | 127.9 (18.4) | 131.2 (16.9) | 135.7 (16.4) |
| Diastolic BP (mmHg) | 81.3 (9.1) | 81.9 (11.4) | 79.6 (10.5) | 82.3 (9.6) |
| Waist (cm) | 105.5 (7.4) | 105.1 (9.8) | 97.9 (8.3) | 107.0 (8.9) |
| Central obesity (%) | 91.9% | 78.5% | 73.6% | 88.0% |
| HbA1c (%) | 5.6 (0.5) | 5.9 (0.4) | 5.8 (0.4) | 5.7 (0.4) |
| Prediabetes (%) | 32.9% | 54.7% | 56.6% | 38.1% |

The 3,682 study participants are 49.2% female and aged 52.8 (SD 8.2) years. Clinical characteristics are well balanced between Intervention and Usual care sites (**Table 6**). The median number of participants recruited at each study sites is 30 (range 13-54). More than 90% of follow-up visits are scheduled to be complete December 2020. Based on follow-up to end 2019, the observed incidence of T2D in the study population is 6.1% per annum. This compares favourably with the incidence predicted in the study protocol and power calculation.

**Table 6.** Characteristics of participants in Active and Usual care sites. P values are adjusted for differences in age, gender and country.

|  | **Active** | **Usual care** | **P** |
| --- | --- | --- | --- |
| N | 1,844 | 1,838 |  |
| Age | 53.3 (8.7) | 53.7 (8.6) | 0.18 |
| Sex (Female %) | 52.1% | 49.3% | 0.09 |
| Never smoked | 86.2% | 86.1% | 0.97 |
| Height | 162.7 (9.7) | 161.9 (9.7) | 0.32 |
| Weight | 80.3 (13.7) | 79.1 (13.5) | 0.31 |
| BMI | 30.3 (4.5) | 30.2 (4.6) | 0.41 |
| Waist | 103.9 (9.3) | 103.8 (9.4) | 0.70 |
| SBP | 130.7 (16.9) | 131.9 (17.1) | 0.51 |
| DBP | 81.0 (10.2) | 81.6 (10.4) | 0.42 |
| HbA1c | 5.76 (0.43) | 5.74 (0.43) | 0.49 |

## Discussion

We describe the rationale, design and implementation of the iHealth-T2D study, a cluster-randomised trial that sets out to determine whether lifestyle intervention delivered by community health workers, to South Asians with central obesity or prediabetes, reduces risk of T2D over three years. Our study is notable for large sample size, multi-country design, and by the use of tools for screening and intervention that have been adapted for low-resource settings.

T2D is a major chronic disease, usually requiring lifelong pharmacological treatment. Having T2D substantially increases a person’s risk of future myocardial infarction, stroke, heart failure, visual impairment, renal impairment, cancer and other long-term conditions.^20^ T2D is thus a leading global cause of morbidity, mortality and healthcare expenditure. The burden of T2D is especially high in South Asia.^4-6^ The World Health Organisation, United Nations and the International Diabetes Federation all recognise that there is an urgent need for action to reverse the current epidemic of T2D in South Asians.

T2D is preventable by lifestyle intervention comprising improved diet, weight loss and increased physical activity.^7,8^ Although many of the studies have been carried out in European populations, lifestyle interventions have been confirmed to reduce risk of T2D in South Asians.^9^ However, the available literature suggests that the benefits of lifestyle intervention may be lower in South Asians. The reasons are not known but may include differences in disease aetiology, limited understanding or compliance with the intervention, reduced availability of alternate healthy foods or facilities for physical activity, social pressures that inhibit behaviour change, or the fidelity of implementation. The evidence for heterogeneity of effect between populations demonstrates the need for specific assessment of the clinical and cost-effectiveness for diabetes interventions in South Asian individuals from the varied ethnic and population subgroups, and in diverse settings. Furthermore, the evidence-based approaches currently described have not been scaled up, and are not routinely available in clinical practice. Obstacles to sustainable implementation include reliance on the oral glucose tolerance test to identify high-risk individuals, as well as the use of dieticians and other qualified healthcare workers for delivery of the intervention. The human and financial resources needed for implementation of these established approaches have are major obstacles to deployment at scale.

On this background we designed the iHealth-T2D study, with the overall goal of advancing the prevention of T2D in South Asian populations. iHealth-T2D was designed to build on previous work, address key limitations in knowledge, and explore more scalable approaches to delivery of lifestyle intervention amongst South Asians. Our study includes three key innovations compared to previous work. First, we use waist circumference and HbA1c for identification of South Asians at increased risk of T2D. Central obesity is widely recognised to be a defining characteristic for susceptibility to T2D amongst South Asians, and can be assessed rapidly, reproducibly and accurately, using low-technology, low-cost approaches, by people with minimal training, and without a qualified healthcare background. Waist circumference is ideally suited to resource poor settings. The choice of HbA1c was informed by our longitudinal population data, which identifies HbA1c as a highly predictive marker of risk for future diabetes in South Asians that can be rapidly measured using point of contact devices, using a single blood test, and without the need for fasting. Together, these characteristics make HbA1c well-suited to population screening in low-resource settings. Second, we use community health workers, combined with group and telephone contacts, for delivery of the intervention with the goal of improving the cost-effectiveness, scalability and sustainability of implementation. Multiple studies from South Asia, and other low-resource settings, support the view that community health workers can provide clinically effective care for chronic diseases, such as hypertension and diabetes.^10-12^ A role in diabetes prevention has not previously been evaluated. The results of the present study are thus anticipated to inform ongoing task-shifting in primary health care systems, in support of improved prevention and control for non-communicable disease. Finally, out study is notable for its multi-country design and large sample size. This will enable investigation of effectiveness on the intervention in key population subgroups, including different cultural, geographic and socio-economic settings. Our results will thus substantially extend knowledge for implementation and effectiveness for diabetes prevention in a range of contexts relevant to South Asian communities.

Our study does also have some limitations. Our recruitment methods did not enable assessment of response rates in South Asia, raising the possibility of responder bias. However, the characteristics of participants are similar to those reported in other studies, arguing against significant population stratification. Our intervention still involves intervention by a health worker, which may represent an obstacle to scale-up. However, we implement task shifting from traditional health care teams, to community health workers, to promote cost-effectiveness and sustainability. We also note that the intensive interventions using peer-support or digital strategies for lifestyle modification have not shown evidence for diabetes prevention in South Asians.^21,22^ Our experiment will thus add to the spectrum of approaches to delivery, and aims to deliver an optimal combination of resource utilisation and clinical effectiveness. We will not be able to collect individual level health expenditure data, which will limit the accuracy of our health economic evaluation. We will mitigate against this through use of curated datasets that we have created for health-related costs across South Asia.

In summary, the iHealth-T2D study of 3,682 South Asians with central obesity or prediabetes will determine whether lifestyle modification delivered by community health workers will reduce risk of T2D over three years. The iHealth-T2D study offers the opportunity to advance understanding of how best to prevent diabetes amongst South Asians, and thereby address a key public health challenge in this major global ethnic group.

## Data Availability

Data will be available to others on completion of the research, by application to the Steering Committee.

## Declarations

### Ethics approval and consent to participate

Ethical approval was obtained from the Institutional Review Board in each participating country and at each research location before the start of the study. Information sheets and consent forms were made available in the major South Asian languages. Multilingual translators were available as required. Each participant provided informed consent. People unwilling or unable to provide consent were excluded from the study.

### India

- Max Healthcare Institutional Ethic Committee (ref: CT/MSSH/SKT-2/ENDO/IEC/14-40, date 22/04/16)
- Indian Council for Medical Research (ICMR, ref: 55/7/Indo-Foreign-Diab/2014-NCD-II, date 08/02/2016)

### Pakistan

- Punjab Institute of Cardiology Ethical Committee (ref: rtpgme-research-047, date 09/04/16)
- Services Institute of Medical Sciences Institutional Review Board (ref: IRB/2016/222/SIMS, date 12/03/16)

### Sri Lanka

- University of Colombo Ethics Review Committee (ref: EC-16-063, date 23/05/16)
- University of Kelaniya Ethics Review Committee (ref: P/62/05/2016, date 11/05/16)

### UK

- West Midlands - Solihull Research Ethics Committee (ref: 16/WM/0171, date 14/04/2016)

## Acknowledgements

We are most grateful to the participants of the iHealth-T2D study, and all the staff members who have taken part in gathering the data of this study.

## Supplementary Online Materials

**S1**. CHW Handbook. Training manual summarising the content and delivery of the lifestyle intervention. Translated versions available through study website.

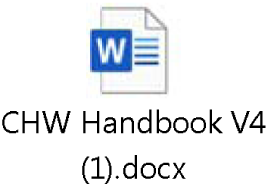

**S2**. CHW Workbook. Case report form / intervention delivery workbook for the lifestyle intervention. Translated versions available through study website.

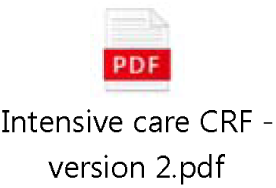

**S3**. Participant Handbook for Lifestyle intervention. Written materials provided to the participant to support the CHW delivered lifestyle intervention. Translated versions available through study website.

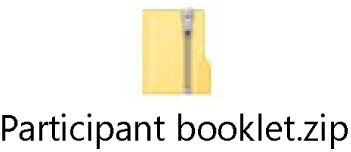

**S4**. Participant Handbook for Usual care. Translated versions available through study website.

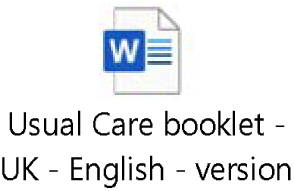

